# A large deletion spanning multiple enhancers near *PITX2* increases primary open-angle glaucoma risk

**DOI:** 10.64898/2026.02.26.25342774

**Authors:** Khaled Said, Ayellet V. Segrè, Janey L. Wiggs, Inas F. Aboobakar

## Abstract

**Importance:** Genome-wide association studies have identified hundreds of common single nucleotide polymorphisms (SNPs) and small insertions/deletions (indels) associated with primary open-angle glaucoma (POAG) risk, though these variants have modest effect sizes and individually may have minor contributions to disease development. As whole-genome sequencing data is becoming more readily available, structural variants and other complex genomic features can be interrogated for contribution to disease risk.

**Objective:** Test the association of structural variants in known glaucoma loci with disease risk.

**Design:** Cross-sectional study.

**Setting:** A multicenter cohort of individuals from the United States who contributed genomic and electronic health record data to the *All of Us* Research Program.

**Participants:** POAG case/control cohorts were generated in the *All of Us* Researcher Workbench using age (>40 for cases, >65 for controls) and ICD 9/10 diagnosis codes.

**Main Outcomes and Measures:** Logistic regression analyses adjusted for age, sex, and the top 10 principal components of ancestry were used to test association of structural variants within 500 kilobases of 309 known open-angle glaucoma risk loci. The significance threshold after Bonferroni correction was set at p<1.6×10^-4^.

**Results:** 516 POAG cases and 18,716 controls of European ancestry from the *All of Us* v8 data release were included in the analysis. Mean age was 77.0 years among cases and 74.7 years among controls. Females comprised 45.7% of cases and 56.5% of controls. An 8,732 base pair deletion upstream of *PITX2* (chr4:110680827-110689558) was associated with 7.3-fold higher odds of POAG (95% confidence interval: 2.9-18.5, p= 2.4×10^-5^, variant carrier frequency= 1.6% in cases and 0.25% in controls). Functional annotation identified multiple enhancers overlapping the deletion, suggesting that this structural variant likely impacts gene regulation and expression.

**Conclusion and Relevance:** Whole genome sequencing data captures rare structural variants with large effect sizes that are missed by conventional SNP and indel genotyping approaches, enabling improved POAG risk stratification. These data also expand the phenotypic spectrum of structural variation in the *PITX2* locus from childhood glaucoma to adult-onset disease, where age at diagnosis and clinical severity may be influenced by the extent of disrupted regulatory elements.

## Introduction

Primary open-angle glaucoma (POAG) is a leading cause of irreversible blindness worldwide, making early detection and timely intervention critical.^1,2^ It is a highly heritable disease, with hundreds of risk loci identified to date.^3-5^ Nearly all single nucleotide polymorphisms (SNPs) and small insertions/deletions (indels) discovered in POAG genome-wide association studies (GWAS) are located in non-coding regions of the genome and have modest effect sizes, suggesting that gene regulation plays a critical role in POAG pathogenesis. While fine-mapping studies have shown that regulation of gene expression or alternative splicing may underlie the causal mechanism for over half of known POAG GWAS loci^6^, many associated variants may also be tagging rare, yet undetected, causal variants with large effect sizes. To date, GWAS have primarily been performed with genotyping arrays, which directly assay only a subset of variants (∼2%) across the genome; the remaining “missing genotypes” are then inferred using ancestry-specific reference panels.^7^ This approach limits discovery to variants that are common in the population, with minimal opportunity to study rare or complex genomic variation that may have larger effects on disease risk. Whole-genome sequencing (WGS) overcomes this limitation, although its higher cost previously limited widespread use.^8^

Structural variants (SVs) are a class of variants that involve large-scale alterations to the genome (>50 base pairs), which includes deletions, duplications, insertions, inversions, translocations, and complex rearrangements.^9^ Due to their size, SVs can have substantial functional consequences, including disruption of coding genes or their *cis*-regulatory elements (ex: promoters, enhancers, and silencers).^10^ Technological advancements, including short- and long-read WGS, have enabled detection of substantially larger numbers of SVs in human genomes than was previously possible with traditional approaches (ex: chromosomal microarrays).^10^ Consequently, SVs are increasingly recognized as important contributors to both Mendelian disorders and complex-inherited diseases.^11^ To date, SVs have been implicated in congenital and syndromic forms of glaucoma^12-16^ as well as familial normal-tension glaucoma.^17^ However, their contribution to POAG risk remains largely unexplored.

The National Institutes of Health *All of Us* (*AoU*) Research Program aims to enroll over one million individuals of diverse backgrounds to improve disease diagnosis, treatment, and prevention.^18^ Data from surveys, WGS, electronic health records (EHRs), physical measurements, and wearable devices are available in the *AoU* Researcher Workbench.^18,19^ To date, short-read WGS data have been released for 414,830 participants (v8), of whom 97,061 have available SV data. In this study, we leverage the *AoU* dataset to test association of both common and rare SVs in known POAG genomic loci with disease risk. With this approach, we have identified a rare deletion spanning multiple enhancer elements near *PITX2* that increases POAG odds by 7.3-fold.

## Methods

Informed consent was obtained for all participants enrolled in *AoU*. The *AoU* IRB follows the regulations and guidance of the National Institutes of Health Office for Human Research Protections for all studies, ensuring that the rights and welfare of research participants are overseen and protected uniformly.^18,19^ The described research adhered to the tenets of the Declaration of Helsinki.

### Case/control definitions

*AoU* participants with available SV data were identified using the cloud-based *AoU* Researcher Workbench. POAG cases were defined as individuals age >40 with an ICD-9/10 diagnosis code for POAG in the electronic health record, but no code for secondary open-angle glaucoma or primary/secondary angle closure glaucoma. Controls were age >65 and did not have any ICD-9/10 diagnosis codes for any form of glaucoma, glaucoma suspect, ocular hypertension, or family history of glaucoma; a higher age threshold was applied to the control cohort to reduce the likelihood of including undiagnosed cases.

### Whole genome sequencing data quality control

The *AoU* WGS protocol has been extensively described previously.^19^ Briefly, short-read WGS was performed on the Illumina NovaSeq 6000 instrument and downstream analyses were performed using the Illumina DRAGEN platform. Quality control (QC) metrics included the following: mean coverage (threshold of ≥30×), genome coverage (threshold of ≥90% at 20×), coverage of hereditary disease risk genes (threshold of ≥95% at 20×), aligned Q30 bases (threshold of ≥8 × 10^10^), contamination (threshold of ≤1%), and concordance to independently processed array data.^19^ Additional sample QC metrics were also applied by *AoU*, including fingerprint concordance, sex concordance, and cross-individual contamination rate.

Genetically-inferred ancestry was derived by *AoU* for all participants with WGS data; the ancestry labels are consistent with those in Genome Aggregation Database (gnomAD), the Human Genome Diversity Project, and the 1000 Genomes Project.^19^ Individuals with kinship scores >0.1 suggesting relatedness (n=30,584), ancestral outliers on genotype principal components analysis (n=987), or sex ploidy mismatch (n=1,476) were also excluded from the analyses described in this manuscript.

### Structural variant calls

An SV callset was released by *AoU* in June 2024 for a subset of participants with available short-read WGS data (n=97,061). The GATK-SV pipeline was used to call SVs of the following types: deletions, duplications, insertions, inversions, translocations, complex events, and unresolved breakends. All samples passed basic quality control and ploidy estimation filters.^20^ Noisy SV calls and potential false positive calls were removed by *AoU*, leaving 1,506,805 SVs across the genome. For the analyses presented in this manuscript, we further excluded SVs with total minor allele count <6, QUAL score <20, >2% missing genotypes, and significant deviations from Hardy-Weinberg equilibrium (p<1×10^-8^) for common SVs (frequency >1% in the dataset).

SVs were then filtered to those located within 500 kilobases (kb) of known open-angle glaucoma risk loci identified in the International Glaucoma Genetics Consortium (IGGC) European GWAS meta-analysis^4^, the IGGC cross-ancestry GWAS meta-analysis^4^, and the multi-trait analysis of GWAS (MTAG) cross-ancestry meta-analysis^5^; a total of 309 unique open-angle glaucoma loci were found among these GWASs. A 500 kb window was chosen given that >90% of functional promoter and enhancer elements are found within this distance from their target gene.^21^ Following filtering, 13,653 SVs were tested for association with POAG, of which 83% were rare in the dataset (frequency <1%).

### Logistic regression analyses

Firth logistic regression was performed to test association of each SV with POAG risk given the smaller sample size for cases compared to controls. Age, sex, and the top 10 genotype principal components of ancestry were included as covariates. Given that SVs within 500 kb of 309 independent open-angle glaucoma risk loci were tested, the significance threshold after Bonferroni correction was set at p<1.6×10^-4^. As an additional sensitivity analysis for the SV passing the significance threshold, logistic regression conditioning on the lead GWAS variant at that locus (rs17527016[C])^5^ was performed to confirm the SV association remained significant and independent of the lead SNP association.

### *In-silico* annotation

To investigate potential functional consequences of the deletion associated with POAG risk (chr4:110,680,827–110,689,558), this region was annotated with candidate *cis*-regulatory elements (ENCODE GRCh38 cCREs)^22^, predicted transcription factor binding motifs (JASPAR 2024)^23^, and DNase hypersensitivity peaks^24^ that represent regions with possible regulatory activity. These annotations were visualized using the UCSC Genome Browser (https://genome.ucsc.edu/). The SV frequency in different ancestral groups was queried using gnomAD SVs v4.1.0.^25^ The SV was also queried in CADD-SV v1.1 to assess its predicted functional impact.^26^

## Results

### Discovery of a rare structural variant in the *PITX2* locus associated with POAG risk in the *All of Us* Research Program

Our analyses were restricted to individuals of European ancestry with available SV data in the *AoU* v8 data release (516 cases/18,716 controls), as no variants met the Bonferroni-corrected significance threshold in other ancestries, likely due to the smaller sample size (15-298 POAG cases in other ancestries). Following variant-level quality control, 13,653 SVs located within 500 kilobases of 309 previously identified open-angle glaucoma risk loci^5^ remained, 83% of which were rare in the *AoU* European case/control dataset (SV frequency <1%). Among these SVs, 33.8% were deletions, 24.3% duplications, 22.2% insertions, 19.1% unresolved breakends, 0.5% complex events, and <0.1% inversions.

The SVs were tested for POAG association using firth logistic regression adjusted for age, sex, and the top 10 genotype principal components to correct for ancestral substructure. Interestingly, an 8,732 base pair deletion (chr4:110,680,827-110,689,558) located 38,821 base pairs upstream of the *PITX2* transcription start site reached Bonferroni-corrected significance (POAG odds ratio (OR) = 7.3, 95% confidence interval (CI): 2.9-18.5, p= 2.41 × 10^-5^) (**Table 1**). All carriers of this deletion were heterozygous, with an allele frequency of 1.6% in POAG cases (n=8) and 0.25% in controls (n=45). To ensure that the SV association with POAG risk was not driven by the lead GWAS variant at this locus (rs17527016, OR= 1.08 in prior cross-ancestry meta-analyses^4,5^), conditional logistic regression was performed by adding the lead GWAS variant as a covariate in the model. This did not alter the magnitude or strength of the SV association, suggesting its effect is independent of the lead GWAS variant (Table 1). Notably, rs17527016[C] was not significantly associated with POAG risk in this cohort, likely due to the smaller sample size, but its effect size estimate was consistent with prior GWAS findings (OR= 1.1, 95% CI: 0.95-1.3, p= 0.19). The SV was also queried in other ancestries in *AoU* (African, Latino/Admixed American, East Asian, and South Asian), though POAG odds were unable to be calculated due to zero counts in either cases and/or controls, depending on ancestry.

**Table 1.**
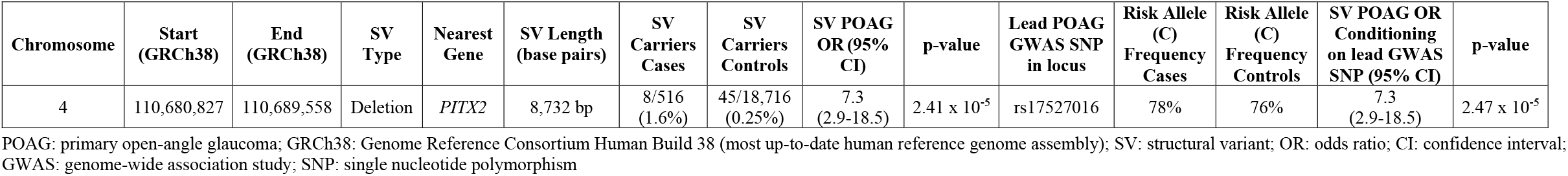
Association of a rare structural variant with POAG in the *All of Us* Research Program.

The novel POAG-associated deletion, which was detected using short-read WGS data, was a high-quality SV call supported by three algorithms (Depth, Manta, and Wham) and four types of evidence (B-allele frequency, paired-end reads, read depth, and split reads). In the gnomAD database, the SV allele frequency is 0.14% in Europeans, 0.02% in Africans, 0.04% in Latino/Admixed Americans, and 0% in East and South Asians.^25^

An additional 265 rare and common SVs reached nominal significance (p<0.05) in the *AoU* European POAG case/control dataset, which are listed in **eTable 1**.

### *In silico* functional annotation reveals multiple enhancer elements spanning the SV region

To assess whether the SV associated with POAG risk in *AoU* is likely to have a functional impact, the variant was queried in the CADD-SV database^26^, where it had a PHRED-scaled score of 14.9 suggestive of moderate deleteriousness. Given that SVs can functionally contribute to disease through disruption of gene regulatory elements (ex: promoters, enhancers, or silencers), we annotated the deletion region (chr4:110,680,827-110,689,558) upstream of *PITX2* with ENCODE candidate *cis*-regulatory elements (cCREs) (**Table 2, Figure 1**). Notably, this region contains four enhancer elements and one chromatin accessibility site that are conserved across mammalian species. Each cCRE also overlaps multiple transcription factor binding motifs and exhibits open chromatin signals on DNase-seq and ATAC-seq indicative of regulatory activity (Table 2). Taken together, these data suggest that the deletion may modify *PITX2* expression levels.

**Table 2.**
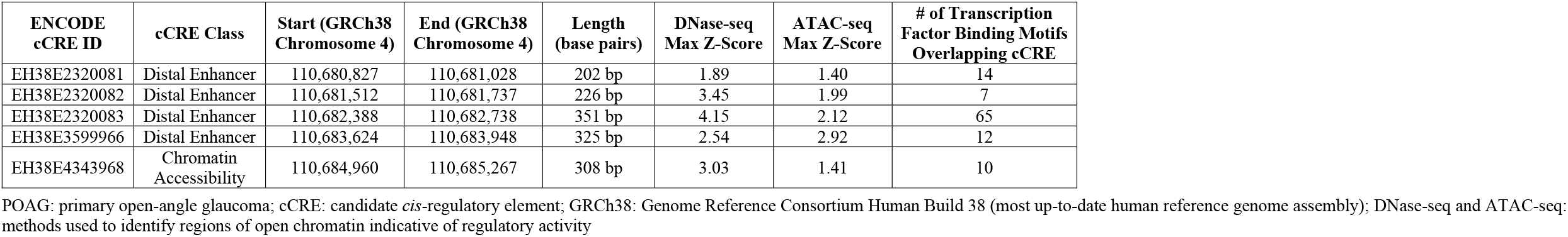
Candidate *cis*-regulatory elements overlapping the POAG-associated deletion (chr4:110,680,827–110,689,558 near *PITX2*).

**Figure 1.**
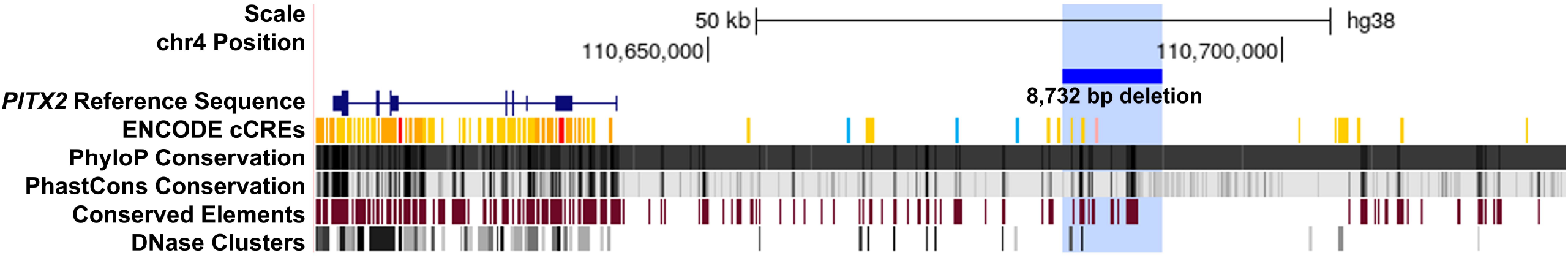
Genomic features spanning the structural variant associated with POAG risk in *All of Us*. A custom track was created in the UCSC genome browser to display the novel 8,732 base pair deletion associated with POAG risk in the *All of Us* Research Program (chr4:110,680,827–110,689,558; blue bar). This region contains multiple candidate *cis*-regulatory elements (cCREs; Table 2) that are conserved across mammalian species, as well as DNase I hypersensitivity clusters that mark active regulatory regions. Of note, *PITX2* is located on the minus strand, so upstream regulatory regions lie at higher genomic coordinates.

## Discussion

To date, POAG genomic discovery has largely been confined to SNPs and small indels captured by conventional genotyping arrays, which typically have modest effects on disease risk and may be genetic markers tagging other causal variants. Fine mapping of risk loci to identify causal variants remains an important unmet need for refining POAG risk prediction models and understanding disease mechanisms. As the cost of WGS has declined, these data are becoming increasingly accessible, enabling interrogation of additional genomic features that may contribute to disease risk. In this study, we leveraged short-read WGS data from the *All of Us* Research Program to test the association of rare and common SVs in known glaucoma risk loci. Using this approach, we identified a rare, putative causal 8,732 base pair deletion spanning multiple enhancers upstream of *PITX2* that increases POAG odds by 7.3-fold.

The *PITX2* gene encodes a homeobox transcription factor that is critical for ocular, craniofacial, and cardiovascular development.^27^ Intriguingly, genetic variation at this locus is associated with a broad phenotypic spectrum: rare *PITX2* coding variants cause congenital and childhood glaucoma in the context of anterior segment dysgenesis and Axenfeld-Rieger syndrome, while a common non-coding SNP identified in GWASs modestly increases POAG risk (OR=1.08).^4,5,28^ WGS studies in families with Axenfeld-Rieger syndrome have also uncovered large deletions of regulatory elements upstream of *PITX2* that segregate with disease.^12-14^ Importantly, functional studies in zebrafish and mouse models show that these SVs reduce *PITX2* expression and recapitulate disease features.^12,14^

The three SVs previously identified in Axenfeld-Rieger families^12-14^ share a 445,359 base pair region of overlap (chr4:110,875,898-111,321,256), which spans a greater number of regulatory elements than the 8,732 base pair deletion associated with POAG risk in *AoU* (chr4:110,680,827-110,689,558) (**Figure 2**). In the CADD-SV database^26^, the SVs linked to early-onset disease have PHRED-scaled scores of 21.5-22.3, indicating greater predicted deleteriousness than the POAG-associated SV, which has a PHRED-scaled score of 14.9. These data suggest that age at diagnosis and clinical severity associated with *PITX2* structural variation exhibits a phenotypic spectrum that is influenced by the extent of disrupted regulatory elements.

**Figure 2.**
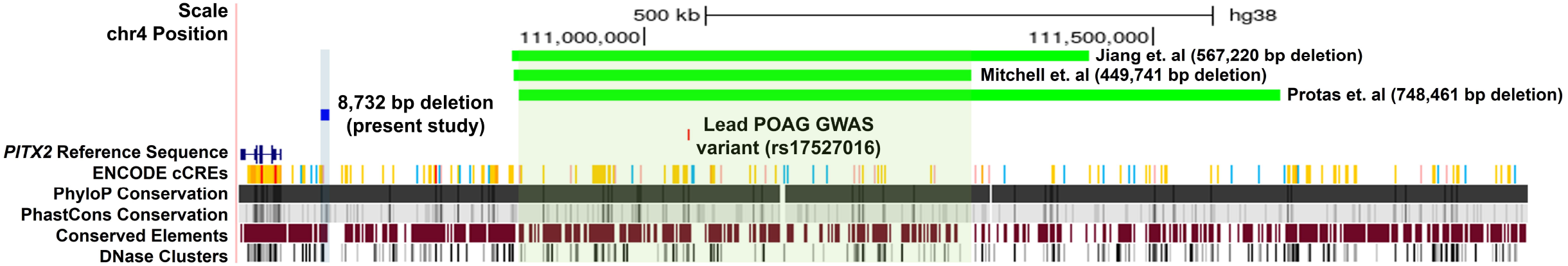
POAG-associated deletion overlaid with previously reported Axenfeld-Rieger deletions and the lead POAG GWAS variant. Custom tracks were created in the UCSC genome browser to display the novel 8,732 base pair deletion associated with POAG risk in the *All of Us* Research Program (chr4:110,680,827–110,689,558; blue bar), previously reported deletions associated with Axenfeld-Rieger syndrome^12-14^ (green bars), and the lead POAG GWAS variant^5^ (chr4:111,042,563; red bar). The three previously reported Axenfeld-Rieger deletions share a 445,359 base pair region of overlap (chr4:110,875,898-111,321,256; highlighted in light green), which contains several conserved candidate *cis*-regulatory elements (cCREs) and DNase I hypersensitivity clusters suggestive of regulatory activity. In contrast, the POAG-associated deletion spans a smaller number of cCREs and DNase I hypersensitivity clusters (highlighted in light blue). Of note, *PITX2* is located on the minus strand, so upstream regulatory regions lie at higher genomic coordinates.

### Limitations

An important limitation of studies utilizing *AoU* is lack of ocular phenotypic data, necessitating use of ICD 9/10 codes from the electronic health record for disease ascertainment. We have previously validated the case/control definitions used in this study, demonstrating strong correlation between effect size estimates for POAG risk loci using our definition with known effect sizes from prior GWAS.^29^ Our study also had limited statistical power with only 516 POAG cases; as *AoU* expands to one million participants, additional POAG-associated SVs may be discovered. Finally, analyses were restricted to individuals of European ancestry due to insufficient sample sizes for other groups, limiting the generalizability of these results to all populations.

## Conclusions

To our knowledge, this is the first study to identify a structural variant that increases risk for POAG, a common, complex-inherited disease. Notably, this SV confers higher disease odds than any previously reported common single nucleotide polymorphism or small insertion/deletion, underscoring the utility of whole-genome sequencing for fine mapping risk loci and refining disease risk stratification. These findings also provide mechanistic insights into the spectrum of *PITX2*-associated glaucoma: SVs spanning a larger number of regulatory elements likely induce greater gene expression changes across tissues and/or cell types, leading to early-onset glaucoma with syndromic features characteristic of Axenfeld-Rieger syndrome, whereas smaller SVs may induce more subtle gene expression changes that manifest as adult-onset POAG.

## Data Availability

The All of Us Research Program does not permit download of individual participant data from the cloud-based server. The code used to perform analyses in the All of Us Researcher Workbench is available upon request.

## Acknowledgements

We gratefully acknowledge *All of Us* Research Program participants for their contributions, without whom this research would not have been possible. Our funding sources include the National Institutes of Health (K23EY035734 to I.F.A., R01EY022305 to J.L.W., R01 EY031820 to J.L.W., R01EY032559 to J.L.W., R01EY031424 to A.V.S., and P30EY014104 to Massachusetts Eye and Ear), Research to Prevent Blindness (Tom Wertheimer Career Development Award in Data Science to I.F.A., Unrestricted Grant to Massachusetts Eye and Ear), and the American Glaucoma Society (MAPS Award and Young Clinician-Scientist Grant to I.F.A.). The sponsor or funding organizations had no role in the design or conduct of this research. The authors have no conflicts of interest to disclose.

## Author contributions

I.F.A. had full access to all the data in the study and takes responsibility for the integrity of the data and the accuracy of the data analysis. **Data curation**: K.S., I.F.A.; **Methodology**: K.S., A.V.S., J.L.W., I.F.A.; **Formal analysis**: I.F.A. **Funding acquisition**: I.F.A.; **Supervision**: I.F.A.; **Writing-original draft**: K.S., I.F.A.; **Writing-review and editing**: K.S., A.V.S., J.L.W., I.F.A.

## Notes

### Competing Interest Statement

The authors have declared no competing interest.

### Author Declarations

Informed consent was obtained for all participants enrolled in the All of Us (AoU) Research Program. The AoU IRB follows the regulations and guidance of the National Institutes of Health Office for Human Research Protections for all studies, ensuring that the rights and welfare of research participants are overseen and protected uniformly. The described research adhered to the tenets of the Declaration of Helsinki.

